# Dengue among febrile patients in Ghana: prevalence and facility-level variation in a multisite surveillance study

**DOI:** 10.64898/2026.11.09.26367667

**Authors:** Francisca Zigah, Adwoa A. Agyei-Nkansah, Emmanuel A. Tagoe, Ivanda A.T. Gyapon, Kofi Bonney, Fred Adomako, Damien T. Punguyire, Joseph A. Oliver-Commey, Richard S. Adzesi, Kwamena W. C. Sagoe, Yaw A. Afrane, Prince J. Pappoe-Ashong, Ernest Kenu, Diederick E. Grobbee, Marianne A. B. van der Sande

## Abstract

**Introduction:** Dengue is under-recognized in Ghana, where laboratory testing is limited. We estimated dengue NS1 and/or IgM positivity among febrile patients and described geographical, temporal, and demographic variation.

**Methodology:** We conducted cross-sectional surveillance among consenting febrile patients aged ≥1 year at six public facilities in Greater Accra, Bono East, and Upper West between August 2023 and July 2025. We randomly selected one region within each of three ecological strata and selected facilities with high febrile-case volumes. We approached and recruited consecutive eligible patients. We tested serum for dengue NS1 antigen and IgM antibody by ELISA; positive NS1 samples underwent RT-PCR and serotyping. We estimated proportions with 95% confidence intervals (CI) and used logistic regression to examine adjusted associations.

**Results:** Among 2,936 participants, 204 were NS1- and/or IgM-positive (6.95%; 95% CI, 6.05-7.93%): 31 were NS1-only, 161 IgM-only, and 12 dual-positive. Prevalence ranged from 3.88% (95% CI, 2.19-6.31%) at Kintampo to 12.32% (9.56-15.55%) at Atebubu, both in Bono East. Adjusted odds increased with age (aOR per five years, 1.09; 95% CI, 1.05-1.14). Eight of 43 NS1-positive samples contained dengue RNA: five DENV-1 from Bono East and three DENV-3 from Upper West. There was no clear rainy-dry season difference.

**Conclusions:** Dengue NS1 and/or IgM were detected among febrile patients at all six facilities, and RT-PCR confirmed circulation of DENV-1 and DENV-3. These findings support including dengue in febrile illness evaluation and strengthening laboratory-supported surveillance in Ghana.

**Author summary:** Dengue is probably overlooked in Ghana because its symptoms resemble malaria and other common causes of fever, while laboratory testing is not routinely available. We studied 2,936 febrile patients who attended six public health facilities across three regions of Ghana. Blood samples were tested for a dengue protein found early in infection and for antibodies that usually appear later. Overall, 204 participants (6.95%; 95% confidence interval, 6.05-7.93%) had evidence of acute or recent dengue infection. Dengue was detected at every participating facility, but proportions varied considerably, even among facilities in the same region. Genetic testing detected dengue virus in eight samples and identified virus types 1 and 3. The chance of a positive result increased with age, although this difference was mainly related to antibody detection rather than the early-infection protein. We did not find a simple difference between rainy and dry seasons. Because the facilities were selected for their high numbers of patients with fever, the findings do not estimate dengue prevalence in the general population or across entire regions. They show that dengue contributes to febrile illness in Ghana and support continued surveillance that reports findings separately for individual facilities and diagnostic markers.

## Introduction

Dengue is a mosquito-borne viral infection caused by four dengue virus serotypes (DENV-1-4), transmitted primarily by Aedes aegypti [1, 2]. Infection may be asymptomatic or present as an acute febrile illness, while fewer patients develop severe disease characterized by plasma leakage, bleeding, or organ impairment [1, 2]. In sub-Saharan Africa, dengue remains under-recognized because laboratory testing and surveillance are limited, and its clinical presentation overlaps with malaria and other common causes of fever [3, 4, 5]. Studies of patients with suspected malaria or undifferentiated fever have identified dengue virus or dengue-reactive test results, supporting the use of laboratory testing to investigate its contribution to febrile illness [6, 7].

Previous reports from Ghana indicated both the presence of competent vectors and dengue virus circulation. Entomological investigations documented *Aedes aegypti* across different ecological settings, with variation in mosquito abundance and breeding habitats [8]. Laboratory experiments demonstrated that some Ghanaian Aedes aegypti colonies can transmit DENV-1 [9]. Earlier serological studies detected dengue-reactive antibodies among malaria-positive children [10], while molecular investigation identified dengue virus among children initially assessed for malaria in Accra [6]. Subsequently, analysis of archived samples from febrile patients collected during 2016-2018 demonstrated dengue seroreactivity [11], while surveillance during 2022-2023 detected DENV-1, DENV-2, and DENV-3 [12]. Ghana subsequently reported a dengue outbreak beginning in the Eastern Region in July 2024 [13]. Therefore, Ghana needs to systematically assess dengue prevalence among patients with febrile illness to guide future surveillance and preparedness.

Comparable evidence from multiple health facilities is needed to describe how frequently dengue is detected among febrile patients and how detection varies across locations, age groups, and time. Regional estimates may conceal differences between facilities serving distinct populations, while temporal comparisons require consistent testing and clearly defined denominators. Diagnostic interpretation also matters: NS1 antigen provides evidence of acute infection, whereas IgM may remain detectable after the acute episode. Combining these tests can identify infection across different presentation times [14, 15]. Applying a common surveillance protocol across facilities can provide useful evidence for planning laboratory testing and monitoring local patterns of dengue detection. We conducted facility-based cross-sectional surveillance among febrile patients attending public health facilities in Ghana. We estimated dengue NS1 and/or IgM positivity by ELISA, described geographical and temporal variation, and examined demographic and household factors associated with a positive result. RT-PCR testing and serotyping of NS1-positive samples provided additional evidence of dengue virus circulation.

## Methods

### Study design and setting

Ghana has 16 administrative regions; all were eligible for selection and were grouped into three broad ecological-zone strata to capture geographical and ecological diversity. We randomly selected one region by ballot from each stratum, selecting the Greater Accra Region (GAR), Bono East Region (BER), and Upper West Region (UWR). Within each region, we purposively selected two public facilities. At study initiation, no officially recognized dengue outbreak was available to guide site selection. We therefore reviewed the regional health directorates’ 2022 facility reports and selected facilities reporting comparatively high total numbers of febrile cases within their regions because they could support sentinel surveillance. The underlying facility-level totals were used operationally for site selection and were not analyzed as study data. Recruitment extended from August 2023 to July 2025, with staggered site periods: LEKMA and Maamobi General hospitals in GAR (July 2024-July 2025), Atebubu and Kintampo Municipal hospitals in BER (September 2023-September 2024), Wa Municipal and Wa West District hospitals in UWR (August 2023-August 2024) (Fig 1).

**Figure 1.**
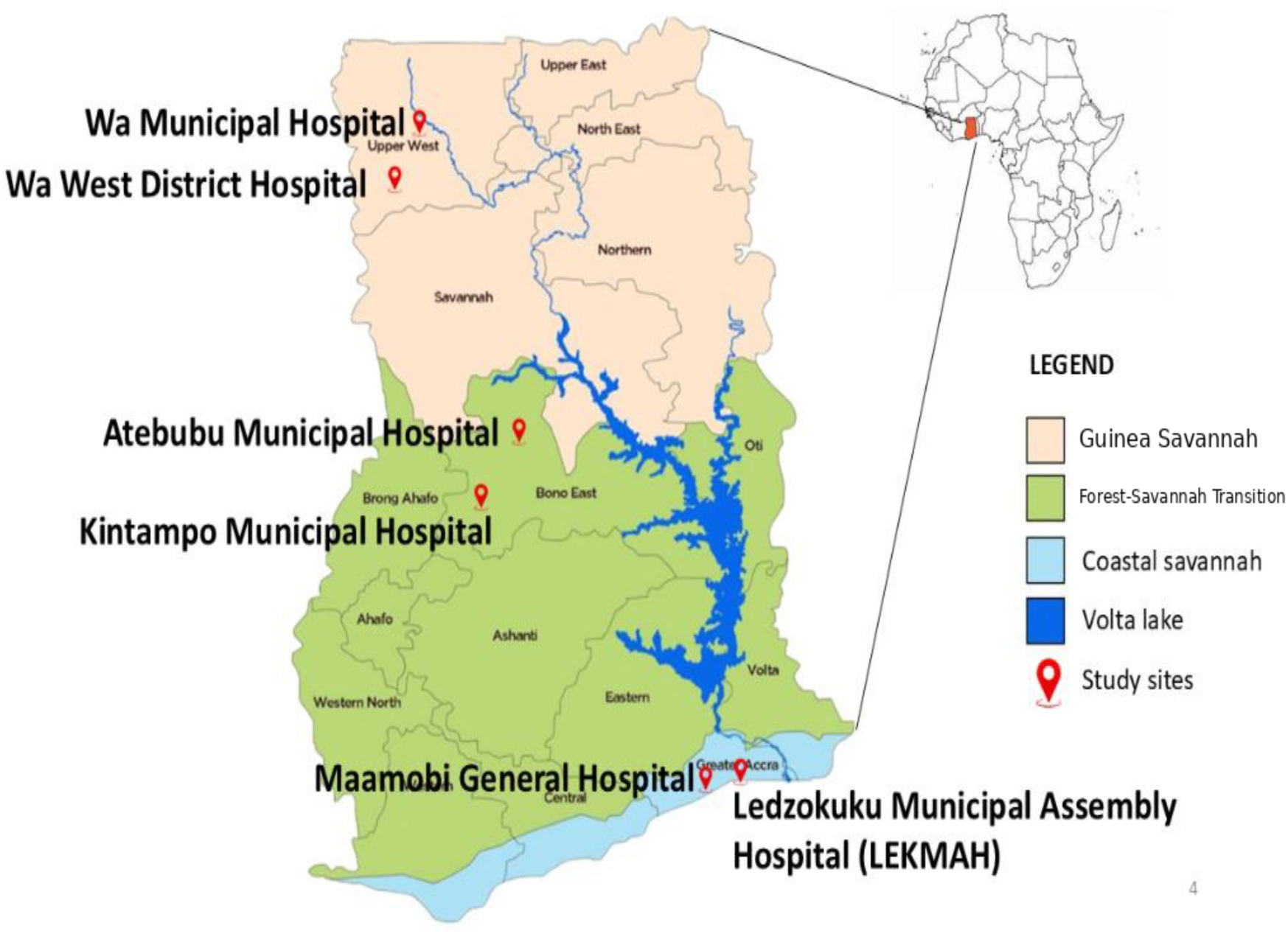
Participating facilities in the ecological context of Ghana (*Map drawn by Yaw Afrane)*

The GAR facilities are located in the Coastal Savannah ecological zone, the BER facilities in the Forest-Savannah Transition zone, and the UWR facilities in the Guinea Savannah zone [16]. For the seasonal analysis, we grouped enrolment months using study-defined regional calendars: rainy months were April-July and September-November in GAR, May-July and September-October in BER, and May-October in UWR; the remaining months were classified as dry. Ecological zone was used to contextualize the facilities and their seasonal patterns, not as an independent analytical exposure. Because each ecological-zone stratum was represented by only one selected region, ecological zone and region were therefore perfectly aligned, and their effects could not be separated.

### Participants and eligibility

Patients aged 1 year old and above were eligible if they had an axillary temperature ≥37.5°C at presentation or reported fever at any time during the preceding seven days, together with at least two compatible symptoms such as chills, headache, retro-orbital pain, myalgia, arthralgia, nausea or vomiting, rash, weakness, malaise, anorexia, lymphadenopathy or bleeding; the seven-day criterion referred to the recency of reported fever, not a maximum total illness duration. Eligible patients were approached in order of presentation from Monday to Friday between 8:00 a.m. and 5:00 p.m. during each site’s active recruitment period. We did not conduct night-time, weekend, or holiday recruitment because attendance during those periods was generally low. The number of eligible patients who declined participation before consent was not recorded.

### Sample size

We calculated the base sample size using a single-population proportion formula with an anticipated prevalence of 5%, 95% confidence interval (CI), and an absolute precision of 1 percentage point. We applied a design effect of 1.5 to account for within-facility clustering and between-facility heterogeneity, yielding a sample size of 2,738 participants. An allowance of approximately 10% for incomplete questionnaires, unusable specimens, or other non-evaluable observations increased the operational recruitment target to 3,042.

### Data and laboratory procedures

Trained clinical staff administered a structured questionnaire covering age, sex, residential setting, water sources and storage, household containers, vector-control practices, reported yellow-fever vaccination, and fever duration. Axillary temperature was measured digitally. We classified reported yellow-fever vaccination as no, yes, or not sure. Fever duration was classified as <3, 3-7, or >7 days. We collected approximately 5 mL of venous blood; we separated serum, stored it temporarily at -20°C, and transported it under temperature-controlled conditions.

### Dengue NS1 and IgM ELISA testing

We analyzed serum samples for the presence of dengue NS1 antigen and anti-dengue IgM using commercial Standard Dengue NS1 Ag ELISA and Standard Dengue IgM ELISA kits (SD Biosensor, Inc., South Korea), respectively, according to the manufacturer’s instructions. Optical densities were measured at 450 nm using a microplate reader, and cut-off values were calculated to determine sample positivity as specified in the kit protocol.

Cut-off optical density = mean calibrator optical density × lot-specific calibration factor. Sample index = sample optical density/cut-off optical density.

We classified index values below 0.9 as negative, 0.9-1.1 as equivocal, and above 1.1 as positive. These correspond to <9, 9-11, and >11 SD Biosensor units, respectively (SD Biosensor units = index × 10). Equivocal results were classified as negative in the primary analysis. The primary outcome was a positive result for NS1 antigen, IgM, or both.

### RNA extraction and RT-PCR

All NS1-positive serum samples underwent reverse-transcription polymerase chain reaction (RT-PCR) testing for dengue virus RNA. Viral RNA was extracted from 140 μL of serum using the QIAamp Viral RNA Mini Kit (Qiagen, Hilden, Germany) and eluted in 60 μL of elution buffer. Dengue virus RNA was detected using the CDC Trioplex real-time RT-PCR assay with AgPath-ID One-Step RT-PCR reagents [17]. Serotypes were identified using the serotype-specific TaqMan real-time RT-PCR assay described by Johnson et al. [18]. Positive and negative controls were included in each run. The molecular results were recorded in a separate laboratory database and were linked to participants using unique study identifiers. Molecular testing was performed at the Noguchi Memorial Institute for Medical Research.

### Ethics approval and consent

The Ghana Health Service Ethics Review Committee approved the study (GHS-ERC 007/12/22). Written informed consent was obtained from adult participants and from parents or guardians of minors; assent was obtained from children where applicable.

### Statistical analysis

We analyzed data using R version 4.5.0. Characteristics were summarized using counts and percentages or medians and interquartile ranges. Dengue NS1 and/or IgM positivity was estimated overall and by region, facility, month, and season, with exact binomial 95% confidence intervals. Monthly comparisons were descriptive because recruitment periods differed and some denominators were small.

The primary logistic regression included age as a continuous variable scaled per five years, sex, facility, residential setting, tap-water use, and empty containers. The primary and all supporting models included 2,936 participants, with no missing observations for the variables included in these models; therefore, no exclusions for missing data or imputation were required. We report adjusted odds ratios (aORs) with Wald 95% confidence intervals. Conventional model-based standard errors were used. Regression estimates were considered exploratory because residual within-facility correlation may affect precision. Supporting analyses examined age groups, a natural cubic spline for age, region in place of facility, season, fever duration, and NS1 and IgM separately. Methods and results are provided in the supplementary material.

## Results

### Participants and dengue test results

Of 3,341 eligible patients who provided study consent, 405 declined venipuncture; 2,936 provided specimens which were analyzed. Median age was 17 years (IQR, 5-33), and 1,664 (56.7%) were female (Table 1). Overall, 204 participants were NS1-and/or IgM-positive (6.95%; 95% CI, 6.05-7.93%), comprising 31 NS1-positive only, 161 IgM-positive only, and 12 NS1-IgM-positive.

**Table 1.** Selected participant characteristics by composite dengue ELISA status.

| Characteristic | Overall (N=2,936) | Negative (N=2,732) | Positive (N=204) |
|---|---|---|---|
| Age, median (IQR), years | 17 (5-33) | 16 (4-32) | 23 (7-42.5) |
| Age 1-4 years | 732 (24.9%) | 694 (25.4%) | 38 (18.6%) |
| Age 5-9 years | 413 (14.1%) | 384 (14.1%) | 29 (14.2%) |
| Age 10-14 years | 241 (8.2%) | 229 (8.4%) | 12 (5.9%) |
| Age 15-19 years | 233 (7.9%) | 224 (8.2%) | 9 (4.4%) |
| Age 20-34 years | 629 (21.4%) | 579 (21.2%) | 50 (24.5%) |
| Age 35-49 years | 339 (11.5%) | 308 (11.3%) | 31 (15.2%) |
| Age ≥50 years | 349 (11.9%) | 314 (11.5%) | 35 (17.2%) |
| Female | 1,664 (56.7%) | 1,546 (56.6%) | 118 (57.8%) |
| Greater Accra | 1,108 (37.7%) | 1,046 (38.3%) | 62 (30.4%) |
| Bono East | 882 (30.0%) | 806 (29.5%) | 76 (37.3%) |
| Upper West | 946 (32.2%) | 880 (32.2%) | 66 (32.4%) |
| Peri-urban residence | 118 (4.0%) | 113 (4.1%) | 5 (2.5%) |
| Rural residence | 1,442 (49.1%) | 1,326 (48.5%) | 116 (56.9%) |
| Urban residence | 1,376 (46.9%) | 1,293 (47.3%) | 83 (40.7%) |
| Uses tap water | 1,873 (63.8%) | 1,758 (64.3%) | 115 (56.4%) |
| Empty containers present | 1,399 (47.6%) | 1,284 (47.0%) | 115 (56.4%) |
| Fever <3 days | 714 (24.3%) | 657 (24.0%) | 57 (27.9%) |
| Fever 3-7 days | 1,495 (50.9%) | 1,383 (50.6%) | 112 (54.9%) |
| Fever >7 days | 727 (24.8%) | 692 (25.3%) | 35 (17.2%) |

Values are n (%) unless otherwise indicated. IQR, interquartile range.

RT-PCR detected dengue RNA in eight of the 43 NS1-positive samples tested (18.60%; 95% CI, 8.39-33.40%): five DENV-1 infections in Bono East and three DENV-3 infections in Upper West. None of the five NS1-positive samples from Greater Accra were RT-PCR-positive (Supplementary Table S1).

### Geographical distribution

Dengue positivity was 5.60% in GAR, 8.62% in BER, and 6.98% in UWR. The largest within-region contrast was between Atebubu (12.32%; 95% CI, 9.56-15.55%) and Kintampo (3.88%; 95% CI, 2.19-6.31%), both in Bono East. Estimates for all six facilities are shown in Table 2.

**Table 2.** Composite dengue ELISA prevalence overall, by region and by participating facility.

| **Location** | **Positive/total** | **Prevalence, % (95% CI)** |
|---|---|---|
| Overall: all facilities | 204/2,936 | 6.95 (6.05-7.93) |
| **Greater Accra** | **62/1,108** | **5.60 (4.32-7.12)** |
| LEKMA Hospital | 37/702 | 5.27 (3.74-7.19) |
| Maamobi General Hospital | 25/406 | 6.16 (4.02-8.96) |
| **Bono East** | **76/882** | **8.62 (6.85-10.67)** |
| Atebubu Municipal Hospital | 61/495 | 12.32 (9.56-15.55) |
| Kintampo Municipal Hospital | 15/387 | 3.88 (2.19-6.31) |
| **Upper West** | **66/946** | **6.98 (5.44-8.79)** |
| Wa Municipal Hospital | 22/351 | 6.27 (3.97-9.34) |
| Wa West District Hospital | 44/595 | 7.39 (5.42-9.80) |

### Factors associated with dengue positivity

The adjusted odds of a positive NS1 and/or IgM result increased with age (aOR per five years, 1.09; 95% CI, 1.05–1.14). Compared with Atebubu, adjusted odds were lower at Kintampo, LEKMA, Maamobi, and Wa West; the comparison with Wa Municipal was inconclusive (aOR, 0.56; 95% CI, 0.29–1.10). There was no clear evidence of an association with sex, residential setting, tap-water use, or empty containers (Table 3).

**Table 3.** Multivariable logistic regression for dengue NS1 and/or IgM positivity with age per five years.

| **Variable** | **aOR** | **95% CI** | **p-value** |
|---|---|---|---|
| Age per five years | 1.09 | 1.05-1.14 | **<0.001** |
| Male versus female | 1.06 | 0.78-1.42 | 0.719 |
| Kintampo versus Atebubu | 0.33 | 0.18-0.60 | **<0.001** |
| LEKMA versus Atebubu | 0.34 | 0.17-0.70 | **0.003** |
| Maamobi versus Atebubu | 0.38 | 0.17-0.85 | **0.018** |
| Wa Municipal versus Atebubu | 0.56 | 0.29-1.10 | **0.091** |
| Wa West versus Atebubu | 0.30 | 0.18-0.50 | **<0.001** |
| Peri-urban versus urban | 0.59 | 0.22-1.63 | 0.311 |
| Rural versus urban | 0.97 | 0.54-1.77 | 0.929 |
| Tap-water use yes versus no | 0.78 | 0.55-1.11 | 0.172 |
| Empty containers present versus absent | 1.38 | 0.98-1.94 | 0.068 |

All listed variables were included in the model. Reference categories: female sex, Atebubu Municipal Hospital, urban residence, no tap-water use, and absence of empty containers. Confidence intervals and p-values use Wald methods. aOR, adjusted odds ratio.

### Temporal variation

Monthly prevalence of dengue test positivity varied within regions (Figure 2; Supplementary Table S5). Recruitment periods differed, and some months included fewer participants, limiting comparisons across regions and time.

In the age-adjusted model, there was no significant association between rainy versus dry season and dengue positivity (aOR, 0.89; 95% CI, 0.65-1.21). Region-specific seasonal estimates are provided in Supplementary Table S3.

**Figure 2.**
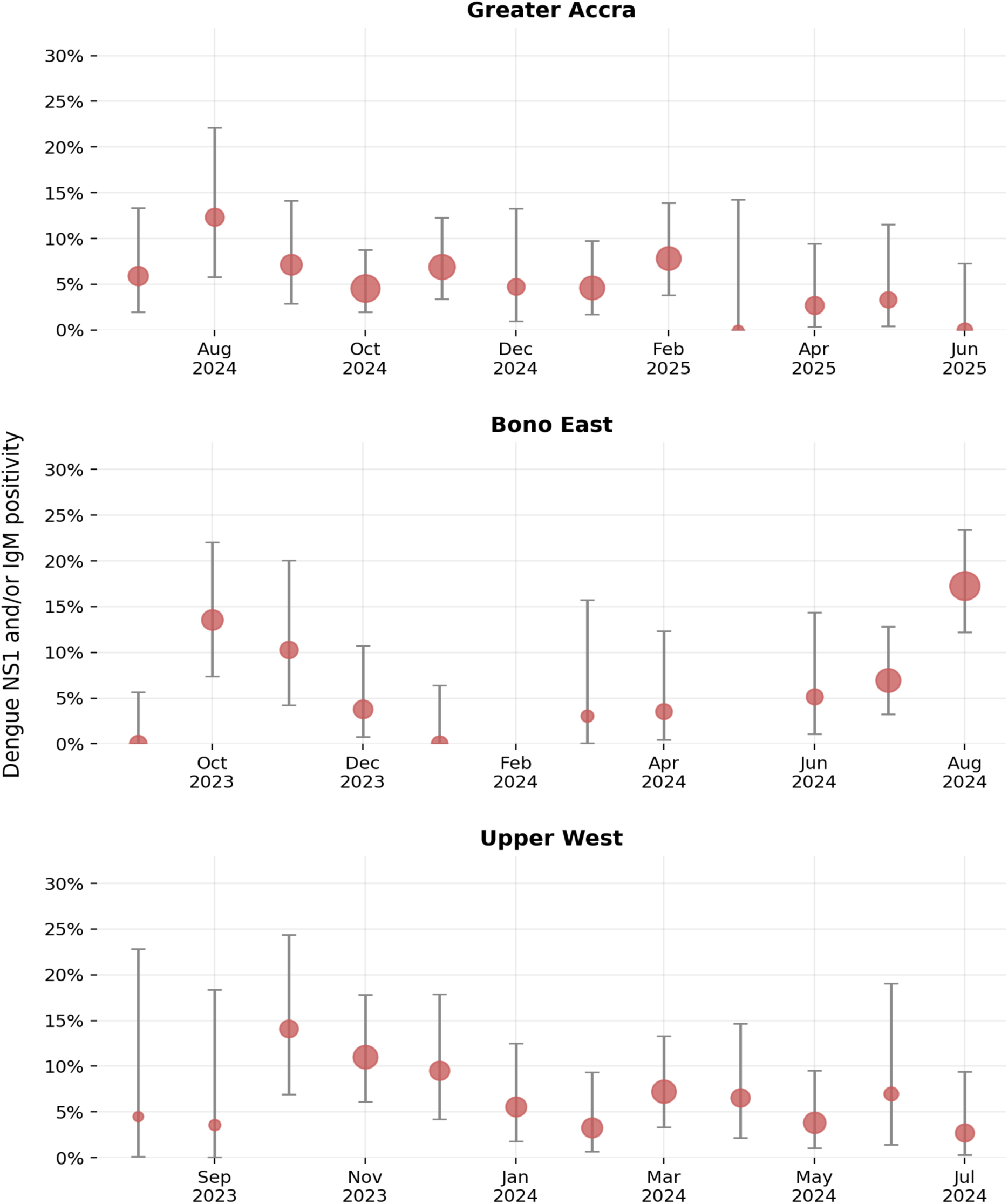
Monthly composite dengue ELISA prevalence by region. Points show monthly proportions and bars show exact 95% confidence intervals; point size reflects the number tested. Months with fewer than 20 participants are omitted from the figure and retained in Supplementary Table S5. Recruitment periods differed by region, so comparisons are descriptive.

## Discussion

This study adds evidence that dengue warrants consideration in the evaluation of febrile illness in Ghana. Detecting NS1 antigen and/or IgM at every participating facility, along with molecular confirmation of DENV-1 and DENV-3, supports strengthening laboratory-supported surveillance beyond individual investigations. The observed differences in prevalence between facilities also show the value of reporting findings at the facility level. These observations provide a basis for improving dengue recognition.

Our findings complement earlier evidence of dengue exposure and viral circulation in Ghana. Stoler et al. detected dengue-reactive antibodies among malaria-positive children, while Amoako et al. subsequently confirmed dengue virus in children initially assessed for malaria [6, 10]. Later serological and molecular studies extended the geographical and temporal evidence [11, 12]. The July 2024 outbreak reported in Ghana further demonstrates the practical relevance of diagnostic preparedness [13]. These observations support maintaining dengue within the differential diagnosis of febrile illness.

Comparisons with other African surveillance studies show why recruitment and diagnostic definitions matter. Lim et al. identified dengue during both outbreak and non-outbreak periods among non-malarial febrile patients in Ouagadougou, using paired serology and selected molecular testing [19]. By contrast, Boyce et al. found few dengue-reactive rapid tests and no PCR-confirmed infections among children with non-malarial fever in western Uganda [7]. A molecular study in Kenya documented substantial dengue detection among febrile children [20]. Although eligibility, age, specimen type, testing strategy, and outbreak context differed, the repeated identification of dengue across African studies supports comparable protocols and clearly defined denominators [21].

The predominance of IgM is central to interpreting our outcome. NS1 and IgM identify overlapping infection windows, and IgM may outlast the presenting episode [14, 15]. Chien et al. detected IgM at prolonged follow-up, with persistence differing between commercial assays [22]. Our combined outcome is therefore an operational measure of NS1 and/or IgM positivity, rather than a precise measure of infections acquired during the reported fever. Because molecular testing was restricted to NS1-positive samples, the molecular findings cannot establish the proportion of all ELISA-positive participants with detectable viral RNA.

Yellow fever exposure and vaccination warrant consideration when interpreting dengue serology, but it should be noted that cross-reactivity may be assay-dependent: Houghton-Triviño et al. observed increased dengue IgM reactivity after yellow fever vaccination, whereas Vázquez et al. found vaccinated participants tested negative with a different dengue-antigen MAC-ELISA [23, 24]. Thus, without confirmatory testing of IgM-only-positive specimens, the contribution of cross-reactivity to dengue NS1 and/or IgM positivity estimates remains uncertain.

The contrast between the two facilities within Bono East illustrates the value of reporting facility-specific findings alongside regional averages. These estimates describe tested patients and cannot directly measure transmission in surrounding communities. Atebubu and Kintampo shared the stated recruitment period, whereas recruitment periods differed across regions, limiting comparisons between regions. Differences in patient populations and attendance may also influence facility estimates. Ghanaian entomological studies document geographical variation in Aedes populations [8, 25], but we did not measure vectors at the participating facilities and cannot attribute the observed differences to mosquito abundance. Facility-specific reporting can identify patterns that require further investigation. The association of increasing prevalence with age mainly involved IgM positivity; the NS1 analysis was less precise. Nevertheless, the clear presence of dengue-reactive antibodies in Ghanaian children and molecularly confirmed infections among Kenyan children show that clinically relevant detection is not restricted to adults [10, 20]. Our findings support clinical awareness across all age groups, without establishing an age threshold for testing.

The absence of a clear rainy–dry season association does not establish an absence of seasonal transmission. Broad seasonal categories may obscure shorter changes in detection, particularly when monthly denominators are small. Surveillance in Burkina Faso identified differences between outbreak and non-outbreak periods [19], supporting interpretation of temporal findings in their epidemiological context. Our staggered recruitment and relatively short observation periods limit inference about recurring seasonal patterns. Repeated surveillance over several years, linked to local rainfall and temperature measurements, would allow a more informative assessment of seasonality.

Several limitations affect the scope of inference. We selected facilities with comparatively high febrile-case volumes, and recruitment was restricted to weekday daytime attendance. The findings therefore cannot be generalized to all health facilities or residents of the participating regions. Consecutive sampling reduced discretionary selection among eligible patients presenting during recruitment hours, but it could not address these broader selection constraints. Pre-consent refusals were not recorded, preventing calculation of an overall participation rate or assessment of the extent of non-participation bias. Single-sample testing limited the ability to distinguish current from recent infection, and self-reported household exposures were subject to misclassification. Adjusted associations remain exploratory because conventional standard errors may not fully account for residual correlation within the six facilities.

Despite these limitations, the harmonized recruitment and testing protocol provides useful guidance for developing dengue surveillance in Ghana. Laboratory-supported sentinel surveillance could improve recognition of dengue among patients with otherwise undifferentiated fever and identify local patterns and risks that require investigation. Reporting antigen, antibody, and molecular findings separately would preserve their distinct interpretations. The findings support expanding access to dengue testing, while further work should assess how to integrate testing into routine febrile-illness care and whether it improves clinical decisions and patient outcomes.

## Conclusion

Dengue NS1 and/or IgM were detected at all six participating facilities, and RT-PCR confirmed DENV-1 and DENV-3 circulation. Consider dengue when evaluating febrile illness in Ghana, with laboratory-supported surveillance to monitor its local distribution.

## Declarations

### Ethics approval and consent

The Ghana Health Service Ethics Review Committee approved the study (GHS-ERC 007/12/22). Written informed consent and, where applicable, parental consent and child assent were obtained.

### Data availability

The de-identified participant-level dataset underlying the findings reported in this manuscript is provided as Supporting Information.

### Author contributions

Conceptualization: FZ, AAN, EAT, FA, DTP, KB, KWCS.

Methodology: FZ, AAN, IATG, FA, DTP, JAO-C.

Funding acquisition: FZ, DEG, MABvdS.

Project administration: FZ, FA, DTP, JAO-C, EK.

Resources: KB, FA, DTP, JAO-C, KWCS, YAA, EK.

Investigation: FZ, EAT, IATG, KB, JAO-C, MABvdS.

Data curation: FZ, IATG, RSA.

Formal analysis: FZ, RSA. Validation: FZ, EAT, MABvdS.

Visualization: FZ, RSA.

Writing-original draft: FZ, AAN.

Writing-review and editing: all authors. All authors approved the final manuscript.

## Acknowledgements

We thank all participants, staff of the six participating hospitals, the Ghana Health Service, Noguchi Memorial Institute for Medical Research, and contributing laboratory staff.

## References

1. World Health Organization. Dengue, https://www.who.int/news-room/fact-sheets/detail/dengue-and-severe-dengue

2. Wilder-Smith A, Ooi EE, Horstick O, Wills B. Dengue. Lancet. 2019;393(10169):350–363. doi:10.1016/S0140-6736(18)32560-1

3. Eltom K, Enan K, El Hussein ARM, Elkhidir IM. Dengue Virus Infection in Sub-Saharan Africa Between 2010 and 2020: A Systematic Review and Meta-Analysis. Front Cell Infect Microbiol. 2021;11:678945. doi:10.3389/fcimb.2021.678945

4. Gainor EM, Harris E, LaBeaud AD. Uncovering the Burden of Dengue in Africa: Considerations on Magnitude, Misdiagnosis, and Ancestry. Viruses. 2022;14(2):233. doi:10.3390/v14020233

5. Stoler J, al Dashti R, Anto F, Fobil JN, Awandare GA. Deconstructing “malaria”: West Africa as the next front for dengue fever surveillance and control. Acta Trop. 2014;134:58–65. doi:10.1016/j.actatropica.2014.02.017

6. Amoako N, Duodu S, Dennis FE, Bonney JHK, Asante KP, Ameh J, et al. Detection of Dengue Virus among Children with Suspected Malaria, Accra, Ghana. Emerg Infect Dis. 2018;24(8):1544–1547. doi:10.3201/eid2408.180341

7. Boyce RM, Collins M, Muhindo R, Nakakande R, Ciccone EJ, Grounds S, et al. Dengue in Western Uganda: a prospective cohort of children presenting with undifferentiated febrile illness. BMC Infect Dis. 2020;20(1):835. doi:10.1186/s12879-020-05568-5

8. Owusu-Asenso CM, Mingle JAA, Weetman D, Afrane YA. Spatiotemporal distribution and insecticide resistance status of Aedes aegypti in Ghana. Parasit Vectors. 2022;15(1):61. doi:10.1186/s13071-022-05179-w

9. Amoa-Bosompem M, Kobayashi D, Itokawa K, Murota K, Faizah AN, Azerigyik FA, et al. Determining vector competence of Aedes aegypti from Ghana in transmitting dengue virus serotypes 1 and 2. Parasit Vectors. 2021;14(1):228. doi:10.1186/s13071-021-04728-z

10. Stoler J, Delimini RK, Bonney JHK, Oduro AR, Owusu-Agyei S, Fobil JN, et al. Evidence of Recent Dengue Exposure Among Malaria Parasite-Positive Children in Three Urban Centers in Ghana. Am J Trop Med Hyg. 2015;92(3):497–500. doi:10.4269/ajtmh.14-0678

11. Pratt D, Takaya H, Abankwa AA, Awuku-Larbi Y, Nyarko S, Agbosu EE, et al. Serologic evidence of dengue and chikungunya among patients with acute febrile illness in Ghana, 2016-2018. J Clin Virol Plus. 2024;4(4):100193. doi:10.1016/j.jcvp.2024.100193

12. Bonney JHK, Pratt D, Ofori M, Hayashi T, Abankwa A, Awuku-Larbi Y, et al. Molecular detection of dengue virus from febrile patients in Ghana. BMC Infect Dis. 2024;24(1):1382. doi:10.1186/s12879-024-10289-0

13. World Health Organization, Ghana Country Office. 2024 annual report: WHO Ghana, https://www.afro.who.int/sites/default/files/2025-06/WHO%20Ghana%202024%20Annual%20Report.pdf

14. Centers for Disease Control and Prevention. Clinical testing guidance for dengue, https://www.cdc.gov/dengue/hcp/diagnosis-testing/

15. Hunsperger EA, Muñoz-Jordán J, Beltran M, Colón C, Carrión J, Vazquez J, et al. Performance of Dengue Diagnostic Tests in a Single-Specimen Diagnostic Algorithm. J Infect Dis. 2016;214(6):836–844. doi:10.1093/infdis/jiw103

16. Yamba EI, Aryee JNA, Quansah E, Davies P, Wemegah CS, Osei MA, et al. Revisiting the agro-climatic zones of Ghana: A re-classification in conformity with climate change and variability. PLOS Clim. 2023;2(1):e0000023. doi:10.1371/journal.pclm.0000023

17. Santiago GA, Vázquez J, Courtney S, Matías KY, Andersen LE, Colón C, et al. Performance of the Trioplex real-time RT-PCR assay for detection of Zika, dengue, and chikungunya viruses. Nat Commun. 2018;9(1):1391. doi:10.1038/s41467-018-03772-1

18. Johnson BW, Russell BJ, Lanciotti RS. Serotype-Specific Detection of Dengue Viruses in a Fourplex Real-Time Reverse Transcriptase PCR Assay. J Clin Microbiol. 2005;43(10):4977–4983. doi:10.1128/JCM.43.10.4977-4983.2005

19. Lim JK, Seydou Y, Carabali M, Barro A, Dahourou DL, Lee KS, et al. Clinical and epidemiologic characteristics associated with dengue during and outside the 2016 outbreak identified in health facility-based surveillance in Ouagadougou, Burkina Faso. PLoS Negl Trop Dis. 2019;13(12):e0007882. doi:10.1371/journal.pntd.0007882

20. Shah MM, Ndenga BA, Mutuku FM, Vu DM, Grossi-Soyster EN, Okuta V, et al. High dengue burden and circulation of 4 virus serotypes among children with undifferentiated fever, Kenya, 2014-2017. Emerg Infect Dis. 2020;26(11):2638–2650. doi:10.3201/eid2611.200960

21. Mwanyika GO, Mboera LEG, Rugarabamu S, Ngingo B, Sindato C, Lutwama JJ, et al. Dengue Virus Infection and Associated Risk Factors in Africa: A Systematic Review and Meta-Analysis. Viruses. 2021;13(4):536. doi:10.3390/v13040536

22. Chien YW, Liu ZH, Tseng FC, Ho TC, Guo HR, Ko NY, et al. Prolonged persistence of IgM against dengue virus detected by commonly used commercial assays. BMC Infect Dis. 2018;18(1):156. doi:10.1186/s12879-018-3058-0

23. Houghton-Triviño N, Montaña D, Castellanos J. Dengue-yellow fever sera cross-reactivity; challenges for diagnosis. Rev Salud Publica (Bogota). 2008;10(2):299–307. doi:10.1590/s0124-00642008000200010

24. Vázquez S, Valdés O, Pupo M, Delgado I, Álvarez M, Pelegrino JL, et al. MAC-ELISA and ELISA inhibition methods for detection of antibodies after yellow fever vaccination. J Virol Methods. 2003;110(2):179–184. doi:10.1016/s0166-0934(03)00128-9

25. Akyea-Bobi NE, Akorli J, Opoku M, Akporh SS, Amlalo GK, Osei JHN, et al. Entomological risk assessment for transmission of arboviral diseases by Aedes mosquitoes in a domestic and forest site in Accra, Ghana. PLoS One. 2023;18(12):e0295390. doi:10.1371/journal.pone.0295390

