## Supplementary Tables for "Dengue among febrile patients in Ghana: prevalence and facility-level variation in a multisite surveillance study"

### **Supplementary material**

#### **Supplementary analytical methods**

Supporting analyses assessed the sensitivity of findings to age specification, geographical adjustment, and diagnostic outcome. Age was categorized as 1–4, 5–9, 10–14, 15–19, 20–34, 35–49, and ≥50 years, with 1–4 years as the reference category. These models retained sex, residential setting, tap-water use, and empty containers, with either facility or region included as a fixed effect (Tables S2 and S6). A natural cubic spline with four degrees of freedom, internal knots at the age quartiles and boundary knots at the minimum and maximum ages was compared with the primary linear-age model using a likelihood-ratio test.

Season was added to the region-adjusted categorical-age model, and positivity was summarized by region and season (Table S3). Fever duration (<3, 3–7, or >7 days) was added separately to examine whether presentation timing affected associations. Separate models examined NS1 and IgM positivity. Because only 43 participants were NS1-positive, the NS1 model included continuous age, sex, region, and fever duration. The IgM model retained the region-adjusted covariates and additionally included reported yellow-fever vaccination (no, yes, or not sure; Table S4).

Monthly positivity was calculated using the number tested in each region and month, with exact binomial 95% confidence intervals; months with fewer than 20 participants were flagged (Table S5). Temporal comparisons were descriptive because recruitment periods differed. Regression estimates used conventional model-based standard errors and Wald confidence intervals; global tests used likelihood-ratio tests. All supporting analyses were exploratory.

#### **Supplementary Result**

There was no clear evidence that the natural cubic spline improved fit compared with linear age (likelihood-ratio p=0.444).

**Supplementary Table S1. Dengue virus serotyping**

| **Region** | **NS1 positive and tested** | **RT-PCR positive** | DENV-1 | **DENV-3** | **RT-PCR positivity** |
| --- | --- | --- | --- | --- | --- |
| Greater Accra | 5 | 0 | 0 | 0 | 0.0% |
| Bono East | 25 | 5 | 5 | 0 | 20.0% |
| Upper West | 13 | 3 | 0 | 3 | 23.1% |
| **Total** | **43** | **8** | **5** | **3** | **18.6%** |

Supplementary Table S2. Region-adjusted sensitivity logistic regression model.

| **Variable** | **aOR** | **95% CI** | **p-value** |
| --- | --- | --- | --- |
| **Age group** | - | - | <0.001 |
| 1-4 years | Reference | - |  |
| 5-9 years | 1.38 | 0.84-2.28 | 0.208 |
| 10-14 years | 0.95 | 0.48-1.85 | 0.870 |
| 15-19 years | 0.85 | 0.40-1.82 | 0.680 |
| 20-34 years | 2.25 | 1.39-3.66 | <0.001 |
| 35-49 years | 2.57 | 1.50-4.40 | <0.001 |
| ≥50 years | 3.04 | 1.77-5.23 | <0.001 |
| **Sex** | - | - | 0.831 |
| Female | Reference | - |  |
| Male | 1.03 | 0.77-1.39 | 0.831 |
| **Region** | - | - | <0.001 |
| Bono East | Reference | - |  |
| Greater Accra | 0.37 | 0.21-0.68 | 0.001 |
| Upper West | 0.49 | 0.32-0.74 | <0.001 |
| **Residential setting** | - | - | 0.711 |
| Urban | Reference | - |  |
| Peri-urban | 0.79 | 0.29-2.16 | 0.650 |
| Rural | 0.82 | 0.50-1.33 | 0.417 |
| **Tap-water use: yes vs no** | 0.84 | 0.60-1.18 | 0.305 |
| **Empty containers: present vs absent** | 1.33 | 0.95-1.87 | 0.094 |

Bono East was the reference region. Wald coefficient intervals and p-values; likelihood-ratio global p-values.

Supplementary Table S3. Region-specific seasonal prevalence.

| **Region** | **Season** | **Positive/total** | **Prevalence, % (95% CI)** |
| --- | --- | --- | --- |
| Bono East | Dry | 47/500 | 9.40 (6.99-12.30) |
| Bono East | Rainy | 29/382 | 7.59 (5.14-10.72) |
| Greater Accra | Dry | 28/418 | 6.70 (4.50-9.54) |
| Greater Accra | Rainy | 34/690 | 4.93 (3.44-6.82) |
| Upper West | Dry | 44/592 | 7.43 (5.45-9.85) |
| Upper West | Rainy | 22/354 | 6.21 (3.94-9.26) |

Rainy versus dry adjusted OR 0.89 (95% CI, 0.65-1.21).

Supplementary Table S4. Selected sensitivity-analysis estimates.

| **Model** | **Contrast** | **aOR** | **95% CI** | **p-value** |
| --- | --- | --- | --- | --- |
| Composite + fever duration | Age 20-34 vs 1-4 years | 2.05 | 1.26-3.34 | 0.004 |
|  | Age 35-49 vs 1-4 years | 2.37 | 1.38-4.07 | 0.002 |
|  | Age ≥50 vs 1-4 years | 2.82 | 1.64-4.86 | <0.001 |
|  | Greater Accra vs Bono East | 0.27 | 0.14-0.51 | <0.001 |
|  | Upper West vs Bono East | 0.48 | 0.31-0.73 | <0.001 |
|  | Fever >7 vs <3 days | 0.49 | 0.31-0.77 | 0.002 |
| NS1 reduced model | Age, per 10 years | 0.91 | 0.74-1.12 | 0.372 |
|  | Greater Accra vs Bono East | 0.12 | 0.04-0.35 | <0.001 |
|  | Upper West vs Bono East | 0.49 | 0.22-1.05 | 0.068 |
|  | Fever >7 vs <3 days | 0.18 | 0.07-0.49 | <0.001 |
| IgM + yellow fever | Age 20-34 vs 1-4 years | 1.85 | 1.08-3.17 | 0.025 |
|  | Age 35-49 vs 1-4 years | 2.34 | 1.29-4.25 | 0.005 |
|  | Age ≥50 vs 1-4 years | 2.81 | 1.54-5.14 | <0.001 |
|  | Greater Accra vs Bono East | 0.31 | 0.15-0.63 | 0.001 |
|  | Upper West vs Bono East | 0.43 | 0.27-0.70 | <0.001 |
|  | Empty containers present | 1.60 | 1.10-2.33 | 0.014 |
|  | YF vaccination: not sure vs no | 1.50 | 0.93-2.41 | 0.095 |
|  | YF vaccination: yes vs no | 0.99 | 0.60-1.64 | 0.968 |

The NS1 model was reduced because only 43 participants were positive and retained continuous age. Global likelihood-ratio p-values: fever duration in composite model 0.0033; region in NS1 model <0.001; fever duration in NS1 model <0.001; yellow-fever vaccination in IgM model 0.047.

Supplementary Table S5. Complete monthly composite dengue ELISA prevalence.

| **Region** | **Month** | **Positive/total** | **Prevalence, % (95% CI)** | **<20 tested** |
| --- | --- | --- | --- | --- |
| Bono East | Sep 2023 | 0/63 | 0.00 (0.00-5.69) | No |
| Bono East | Oct 2023 | 13/96 | 13.54 (7.41-22.04) | No |
| Bono East | Nov 2023 | 7/68 | 10.29 (4.24-20.07) | No |
| Bono East | Dec 2023 | 3/79 | 3.80 (0.79-10.70) | No |
| Bono East | Jan 2024 | 0/56 | 0.00 (0.00-6.38) | No |
| Bono East | Feb 2024 | 1/17 | 5.88 (0.15-28.69) | Yes |
| Bono East | Mar 2024 | 1/33 | 3.03 (0.08-15.76) | No |
| Bono East | Apr 2024 | 2/56 | 3.57 (0.44-12.31) | No |
| Bono East | May 2024 | 3/17 | 17.65 (3.80-43.43) | Yes |
| Bono East | Jun 2024 | 3/58 | 5.17 (1.08-14.38) | No |
| Bono East | Jul 2024 | 9/129 | 6.98 (3.24-12.83) | No |
| Bono East | Aug 2024 | 33/191 | 17.28 (12.20-23.40) | No |
| Bono East | Sep 2024 | 1/19 | 5.26 (0.13-26.03) | Yes |
| Greater Accra | Jul 2024 | 5/84 | 5.95 (1.96-13.35) | No |
| Greater Accra | Aug 2024 | 9/73 | 12.33 (5.80-22.12) | No |
| Greater Accra | Sep 2024 | 7/98 | 7.14 (2.92-14.16) | No |
| Greater Accra | Oct 2024 | 8/176 | 4.55 (1.98-8.76) | No |
| Greater Accra | Nov 2024 | 10/145 | 6.90 (3.36-12.32) | No |
| Greater Accra | Dec 2024 | 3/63 | 4.76 (0.99-13.29) | No |
| Greater Accra | Jan 2025 | 6/130 | 4.62 (1.71-9.78) | No |
| Greater Accra | Feb 2025 | 10/128 | 7.81 (3.81-13.90) | No |
| Greater Accra | Mar 2025 | 0/24 | 0.00 (0.00-14.25) | No |
| Greater Accra | Apr 2025 | 2/74 | 2.70 (0.33-9.42) | No |
| Greater Accra | May 2025 | 2/60 | 3.33 (0.41-11.53) | No |
| Greater Accra | Jun 2025 | 0/49 | 0.00 (0.00-7.25) | No |
| Greater Accra | Jul 2025 | 0/4 | 0.00 (0.00-60.24) | Yes |
| Upper West | Aug 2023 | 1/22 | 4.55 (0.12-22.84) | No |
| Upper West | Sep 2023 | 1/28 | 3.57 (0.09-18.35) | No |
| Upper West | Oct 2023 | 10/71 | 14.08 (6.97-24.38) | No |
| Upper West | Nov 2023 | 14/127 | 11.02 (6.16-17.80) | No |
| Upper West | Dec 2023 | 8/84 | 9.52 (4.20-17.91) | No |
| Upper West | Jan 2024 | 5/90 | 5.56 (1.83-12.49) | No |
| Upper West | Feb 2024 | 3/91 | 3.30 (0.69-9.33) | No |
| Upper West | Mar 2024 | 9/124 | 7.26 (3.37-13.33) | No |
| Upper West | Apr 2024 | 5/76 | 6.58 (2.17-14.69) | No |
| Upper West | May 2024 | 4/104 | 3.85 (1.06-9.56) | No |
| Upper West | Jun 2024 | 3/43 | 6.98 (1.46-19.06) | No |
| Upper West | Jul 2024 | 2/74 | 2.70 (0.33-9.42) | No |
| Upper West | Aug 2024 | 1/12 | 8.33 (0.21-38.48) | Yes |

Exact Clopper-Pearson confidence intervals are shown. Months with fewer than 20 participants were retained here but omitted from the principal monthly figure.

Supplementary Table S6. Facility-adjusted model with categorical age.

| **Variable** | **aOR** | **95% CI** | **p-value** |
| --- | --- | --- | --- |
| Age group (global) | - | - | **<0.001** |
| 1-4 years | Reference | - |  |
| 5-9 years | 1.37 | 0.83-2.28 | 0.220 |
| 10-14 years | 0.85 | 0.43-1.67 | 0.636 |
| 15-19 years | 0.78 | 0.37-1.68 | 0.532 |
| 20-34 years | 2.18 | 1.32-3.58 | **0.002** |
| 35-49 years | 2.49 | 1.43-4.32 | **0.001** |
| ≥50 years | 3.04 | 1.74-5.31 | **<0.001** |
| Sex: male vs female | 1.08 | 0.80-1.46 | 0.618 |
| Facility (global) | - | - | **<0.001** |
| Atebubu | Reference | - |  |
| Kintampo | 0.33 | 0.18-0.60 | **<0.001** |
| LEKMA | 0.31 | 0.15-0.64 | **0.002** |
| Maamobi | 0.34 | 0.15-0.76 | **0.009** |
| Wa Municipal | 0.58 | 0.30-1.13 | 0.108 |
| Wa West | 0.28 | 0.17-0.47 | **<0.001** |
| Residence (global) | - | - | 0.550 |
| Urban | Reference | - |  |
| Peri-urban | 0.58 | 0.21-1.61 | 0.299 |
| Rural | 0.97 | 0.53-1.77 | 0.928 |
| Tap-water use: yes vs no | 0.77 | 0.54-1.10 | 0.149 |
| Empty containers: present vs absent | 1.37 | 0.97-1.94 | 0.074 |
